# Rural Disadvantage in Glioblastoma Concentrates in the Early Postoperative Period: A Single-Center, Treatment-Standardized Cohort Study

**DOI:** 10.64898/2026.01.06.26343534

**Authors:** Matthew Love, Danny Toon, Abby Mocherniak, Sarah Asselin, Kaytlin Andrews, Alyson Mahar, Shervin Taslimi, James Purzner, Katie Goldie, Teresa Purzner

## Abstract

**Background:** Glioblastoma (GB) is the most common and most aggressive malignant primary brain tumor, with 5-year survival rates around 5%. While treatment-related factors significantly impact outcomes, the influence of sociodemographic variables remains unclear, with prior studies showing inconsistent findings. These inconsistencies may stem from underrepresentation of rural patients within national databases and from comparison across centers with variable treatment practices.

**Objective:** To evaluate whether population size, geographic distance, and regional-average income influence short- and long-term survival among GB patients when treatment delivery is uniform.

**Methods:** We analyzed 248 patients who underwent surgical resection at a single publicly funded tertiary neurosurgical center serving all of Southeastern Ontario (2017-2023). Multivariable logistic regression assessed 90-day mortality, while Cox proportional hazards models with time-varying treatment covariates evaluated overall survival (OS).

**Results:** Among 248 patients, 71.8% resided in communities with <50,000 residents. Residing in areas with ≥50,000 residents was associated with 63% lower odds of 90-day mortality (OR 0.37; 95% CI: 0.17-0.81). Each 10-mile increase in distance was associated with 5% increased odds of 90-day mortality (OR 1.05; 95% CI: 1.01-1.10). Regional-average income showed no association with 90-day mortality (OR 0.96; 95% CI: 0.51-1.81). None of these variables significantly affected OS in multivariable models. Conclusions: When treatment practices are uniform, population size and geographic distance independently influence early post-operative mortality but not long-term survival. These findings suggest that improving equity in GB care requires targeted interventions during the critical first 90 days post-surgery, extending beyond geographic access alone to address challenges inherent to low-population-density communities.

**Key Points:**

- Rural residence and greater distance from tertiary care independently increase 90-day mortality after glioblastoma surgery, even when treatment practices are uniform, suggesting distinct mechanisms requiring separate interventions.
- These sociodemographic factors do not affect overall survival when treatment access is equalized, indicating vulnerability occurs specifically during the early post-operative period rather than throughout the disease course.
- Regional-average income did not influence survival within our universal healthcare system, demonstrating that socioeconomic disparities in GB outcomes may be modifiable through system-level policies ensuring equitable treatment access.

**Importance of Study:** Persistent outcome disparities between rural and urban glioblastoma patients have long been attributed to unequal access to oncologic therapy. By studying a cohort where all patients received care at the same tertiary center with uniform treatment practices, this study isolates the independent effects of geography from treatment variation. Our findings reveal that population size and distance primarily affect 90-day post-operative mortality rather than long-term survival, fundamentally reframing the mechanism through which geography influences these outcomes. This challenges the assumption that simply bringing care closer to rural communities will eliminate disparities. Instead, health systems must develop comprehensive interventions targeting the critical early post-operative period, including enhanced care coordination, proactive follow-up systems, and community-based support programs tailored to rural populations. The null effect of income in our publicly funded system provides compelling evidence that socioeconomic disparities observed in private healthcare systems are modifiable through universal coverage policies. This methodological approach, combining time-varying treatment analysis, uniform care delivery, and representative sampling of underserved populations, offers a replicable framework for investigating disparities in other cancer types and healthcare contexts.

## Introduction

Glioblastomas (GB) are both the most common^1^ and most aggressive malignant brain tumor, having a 5-year survival rate of approximately 5%^2^. Current standard of care involves safe maximal surgical resection, followed by concurrent radiation therapy (RAD) and chemotherapy (CTX)^2^. While the median overall survival (OS) for GB is approximately 15 months, this outcome is typically achieved by patients who receive standard-of-care treatment, such as the STUPP protocol. Survival is markedly shorter for those who do not undergo surgery or adjuvant treatment^3–7^. However, not all patients have equal access to guideline-concordant care, and these persistent disparities represent a key mechanism by which sociodemographic factors influence both quality of care and short- and long-term survival outcomes.

Clinical and treatment-related factors such as age, molecular markers, extent of resection, and pre- or post-surgical functional status have a well-described impact on short- and long-term survival in patients diagnosed with GBs^8–11^, as do certain sociodemographic variables such as ethnicity and sex^12,13^. However, the influence of other sociodemographic variables on GB outcomes, such as regional-level income^3,7,14–23^, population size^3,5,7,24–26^, and distance to tertiary care^6,24,27,28^, remains unclear with studies reporting mixed findings. These inconsistencies may arise from limitations in federal patient data repositories such as the Surveillance, Epidemiology, End-results (SEER) and the National Cancer Database (NCBD), which often underrepresent rural patients. For instance, prior NCBD studies examining the impact of rurality on GB outcomes used a study cohort where rural patients comprised between 2%-5%^5,24^, despite 19% of the United States population residing in rural areas^29^. Consequently, these studies may not fully capture the rural patient experience, including only those who reached commission or cancer-accredited centers that report to the NCDB and potentially excluding patients facing barriers to accessing tertiary care. Furthermore, both SEER and NCDB have faced criticism for comparing patients across vastly different geographic regions^15,30^ with variable treatment practices between neuro-oncology centers, making it challenging to discern whether observed differences in outcomes are due to sociodemographic disadvantages or from variations in care.

To address these limitations, our study investigates differences in 90-day mortality and OS among patients diagnosed with a GB based on population size of resident community, distance to tertiary care, and regional average-income in Southeastern Ontario, Canada. SEO provides an ideal setting to examine how rurality influences survival. It is home to Ontario’s largest rural population (55.8%) as well as several geographically dispersed urban centers, and disproportionately high poverty rates^31^, yet all patients within this region receive brain tumor care from a single, publicly funded neurosurgical and cancer centre, Kingston Health Sciences Centre (KHSC), ensuring uniformity in treatment practices across populations. This unique setting allows us to isolate the effects of sociodemographic factors from treatment variation, providing critical insight to inform equity-focused interventions in geographically dispersed regions.

## Methods

### Data collection

Surgical pathology was collected on all patients who underwent brain tumor surgery between April 1^st^, 2017, and March 31^st^, 2023, at KHSC. To reflect the 2021 CNS WHO classification, our cohort contained only IDH WT GBs, including high-grade histological variants such as gliosarcomas and giant-cell GB tumors with appropriate histological features (microvascular proliferation, necrosis, CDKN2A/B homozygous deletions, TERT mutant, EGFR amplification)^32,33^. Surgical pathology, date of birth (DOB), biological sex, fiscal year of diagnosis, date of operation, MGMT promoter methylation status and postoperative Karnofsky performance score (KPS) were collected on the patient chart system (PCS) affiliated with KHSC from oncology consult notes. Notably, postoperative KPS was used throughout this study, where a score of <70 represented loss of independence and a score of ≥70 indicated functional independence. Date of death (DOD) was determined using PCS death files and, when possible, cross-referenced with online obituaries. Age was calculated as years between DOB and operation. Age was categorized as <65 or ≥65 years for descriptive purposes, and as a continuous variable for regression analysis. Survival was confirmed through clinical appointments and treatment consults up until March 31^st^, 2024, thereby ensuring a minimum of 1 year follow-up for all patients in the cohort. For patients who received multiple operations, survival was calculated based on the time between the first surgery and DOD.

Geographic information including home address, residing town, residing county, and postal code were obtained via PCS for all patients who met the inclusion criteria. The census subdivision associated with patient postal codes was determined using Statistics Canada 2021 census profile table^34^. When postal codes corresponded with more than one census subdivision, the subdivision in closer geographic proximity to the patient address was used. Subdivisions with <50,000 residents were considered lower-populated areas, consistent with provincially and federally recognized definitions of rurality in Canada and the United States^29,35–37^. Proximity to KHSC was determined via the shortest route by land between the patient’s address and the tertiary care centre. As PCS data did not include individual patient income data, regional-average income was used as a proxy and measured using census subdivision average income from Statistics Canada 2021 census profile data^3,24^. Data for both distance to tertiary care and regional-average income were represented as continuous variables, given the absence of consistent federally recognized definitions of remote distance and low regional-average income.

Receipt of post-operative treatment was verified through both RAD and CTX patient appointment histories and oncologist consult notes documented in PCS. The commencement date for RAD was identified as the first appointment starting a series of consecutive RAD appointments (typically pattern of 5 consecutive appointments followed by 2 days off) and validated through RAD consults and completion records. Time to RAD was calculated as the interval between the date of surgical operation (day 0) and the RAD commencement date. For CTX, where the alkylating agent Temozolomide (TMZ) is prescribed during the initial medical oncology appointment, the commencement date was defined as the first medical oncology treatment appointment and confirmed through oncology consult notes. Time to CTX was defined similarly, as the interval between the surgical operation and CTX initiation. If a patient was deceased, then OS was defined as the interval between the surgical operation and DOD. However, if the patient was reported alive prior to the end of follow-up period, then OS was defined as the interval between the surgical operation date and the end of the follow up period. Vital status was assessed up to March 31^st^, 2024, using PCS death files, consult notes, and online obituaries where patient identifiers were cross-referenced to ensure accuracy. For the 90-day mortality outcome, vital status was assessed at 90 days post-surgery; patients who died within this timeframe were classified as deceased, while those alive at 90 days were classified as alive.

### Statistical Analysis

Frequencies of all categorical variables were examined and then stratified by population size, distance to tertiary care, and regional-average income categories. Differences in covariate distributions across population size categories were assessed using Chi-square tests, while differences in covariate-level means for distance to tertiary care and regional average income were evaluated using Mann-Whitney U/Kruskal-Wallis tests^38^. Descriptive statistics were used for all continuous variables, including mean (restricted mean for survival outcomes) and standard deviation. Restricted mean was calculated as the area under the Kaplan-Meier survival curve up to the maximum observed follow-up time (τ =2246 days). Age, distance, and regional-average income were analyzed as continuous variables and were rescaled to reflect effects per 10 years, 10 miles, and $10,000 CAD, respectively, for presentation purposes.

Univariate logistic regression was performed for each variable to estimate the odds ratio (OR) and corresponding 95% confidence interval (C.I.) that assessed the odds associated with 90-day mortality for descriptive purposes. Known confounders (based on literary merit) including fiscal year, MGMT status, age, and KPS were included in all multivariable logistic regression models. To evaluate the adjusted association between each sociodemographic factor of interest (population size, distance to tertiary care, and regional-average income) with 90-day mortality, three separate models were constructed: one including population size, one including distance to tertiary care, and one including regional-average income, each adjusted for identified confounders^39^. However, RAD and CTX were not included in the models due to concerns regarding immortal-time bias^40^. The linearity assumption for continuous variables was checked via the plot smoothed function. Multicollinearity was checked for all variables in the multivariable logistic regression model via variance inflation factor (VIF). *Post-hoc* interaction models were conducted for the sociodemographic variables of interest that demonstrated significance in their respective adjusted multivariable logistic regression model, by creating product terms and evaluating their statistical significance^41,42^.

Univariable Cox proportional hazard models were used to examine the relative rate of mortality associated with the relevant covariates and sociodemographic variables, providing descriptive insight into unadjusted relationships. Hazard ratios (HR) and 95% C.I. were calculated for each independent variable. Adjusted associations between overall mortality hazard and population size, distance to tertiary care, and regional-average income were estimated using separate multivariable Cox proportional hazard models. To accommodate immortal time bias stemming from including RAD and CTX in each model, they were measured as time-dependent covariates. We employed a data restructuring approach that involved splitting each patient’s follow-up time into multiple intervals at designated change points including the commencement of RAD, the commencement of CTX, and the DOD^40,43,44^. Time-to-treatment variables were collected to enable derivation of the time-varying RAD and CTX exposures and were not analyzed as independent variables. Multicollinearity for variables in the Cox proportional hazard model, and proportional hazard assumptions were checked. Violations of multicollinearity led to removal of affected variables in their respective Cox proportional hazard models. Violations of proportional hazards were addressed by using stratification within each of the three cox models, giving the affected variables separate baseline hazards^45^. *Post-hoc* interactions were also planned (again, by creating product terms) based on whether any of population, distance to tertiary care, and regional-average income were deemed significant in their respective adjusted multivariable cox proportional hazard model.

An additional category for missing data was created when the missing data represented over 5% of the variable. We used R version 4.2.1 (2022-06-23) for all analyses. The data extraction was collected using IBM SPSS Statistics (version 29.0.2) and was imported to R studio (version 2024.04.1+748) for statistical analysis and graphics but was edited using Adobe Illustrator 2023 (Version 27.8).

## Results

### Patient Demographics

In total, 1438 brain tumor surgeries were performed at KHSC between April 1st, 2017, and March 31st, 2023, with corresponding surgical pathology reports. Of these, 1163 did not meet histological inclusion criteria, 25 were removed as they represented disease recurrence within the cohort, and an additional 2 patients were lost to follow-up (Figure 1). The final cohort of 248 patients were distributed across all 6 years, with lowest frequency in 2018-2019 (12.5%) and highest in 2020-2021 (22.2%) (Table 1). 55% of the cohort were ≥65 years of age, and 62% of the cohort were male. Over 95% of the diagnoses were GB tumors, with the remaining ∼5% representing other histologic variants of GB. Approximately 41% of the cohort displayed MGMT methylation and over 50% of the cohort had a KPS <70 while 43.6% with KPS ≥70.

**FIGURE 1.**
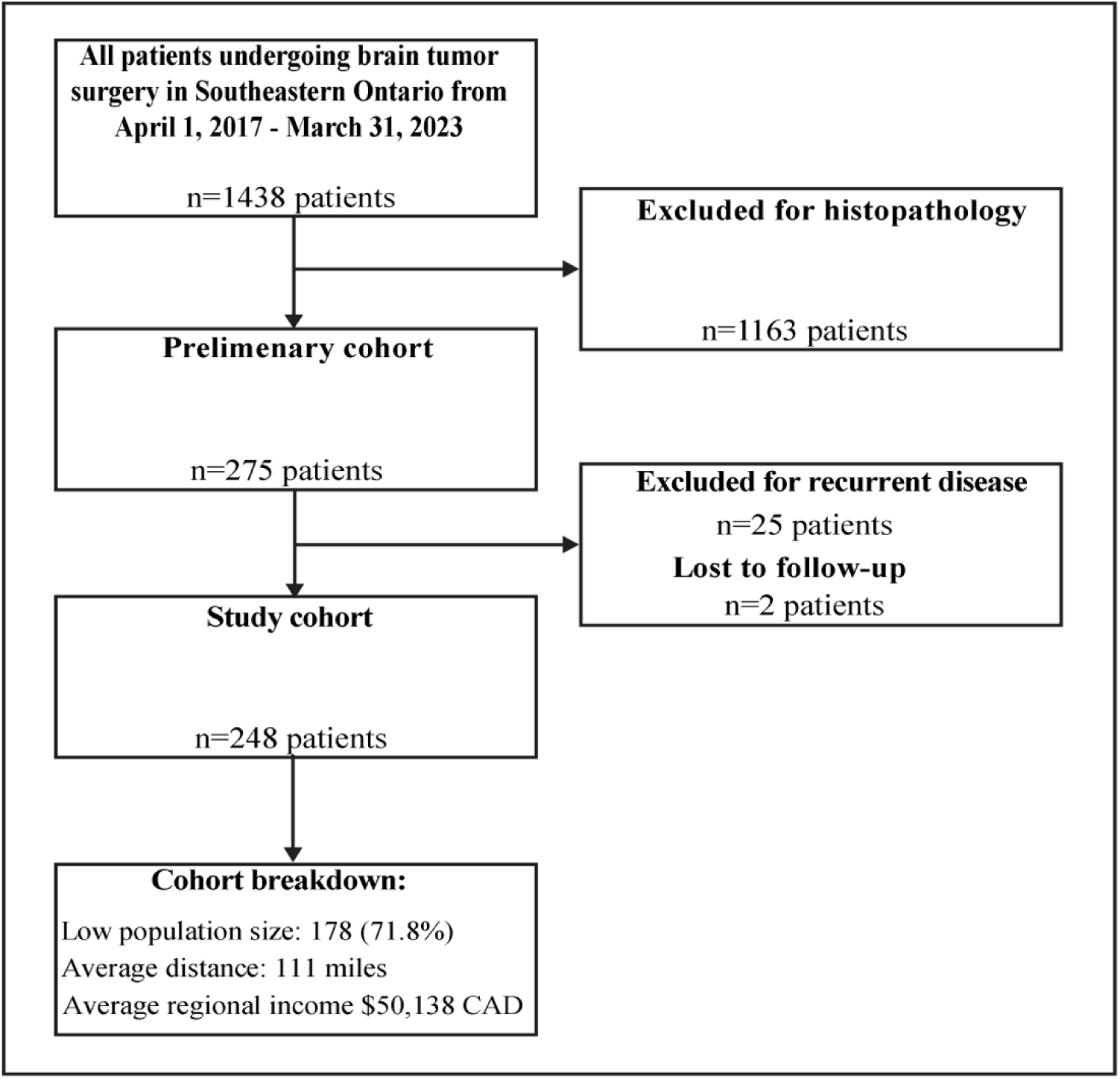
Flowchart of Patient Cohort Selection: Identification of patients who were diagnosed with a glioblastoma and received surgical intervention at Kingston Health Sciences Centre (April 1, 2017 - March 31, 2023).

**TABLE 1.**
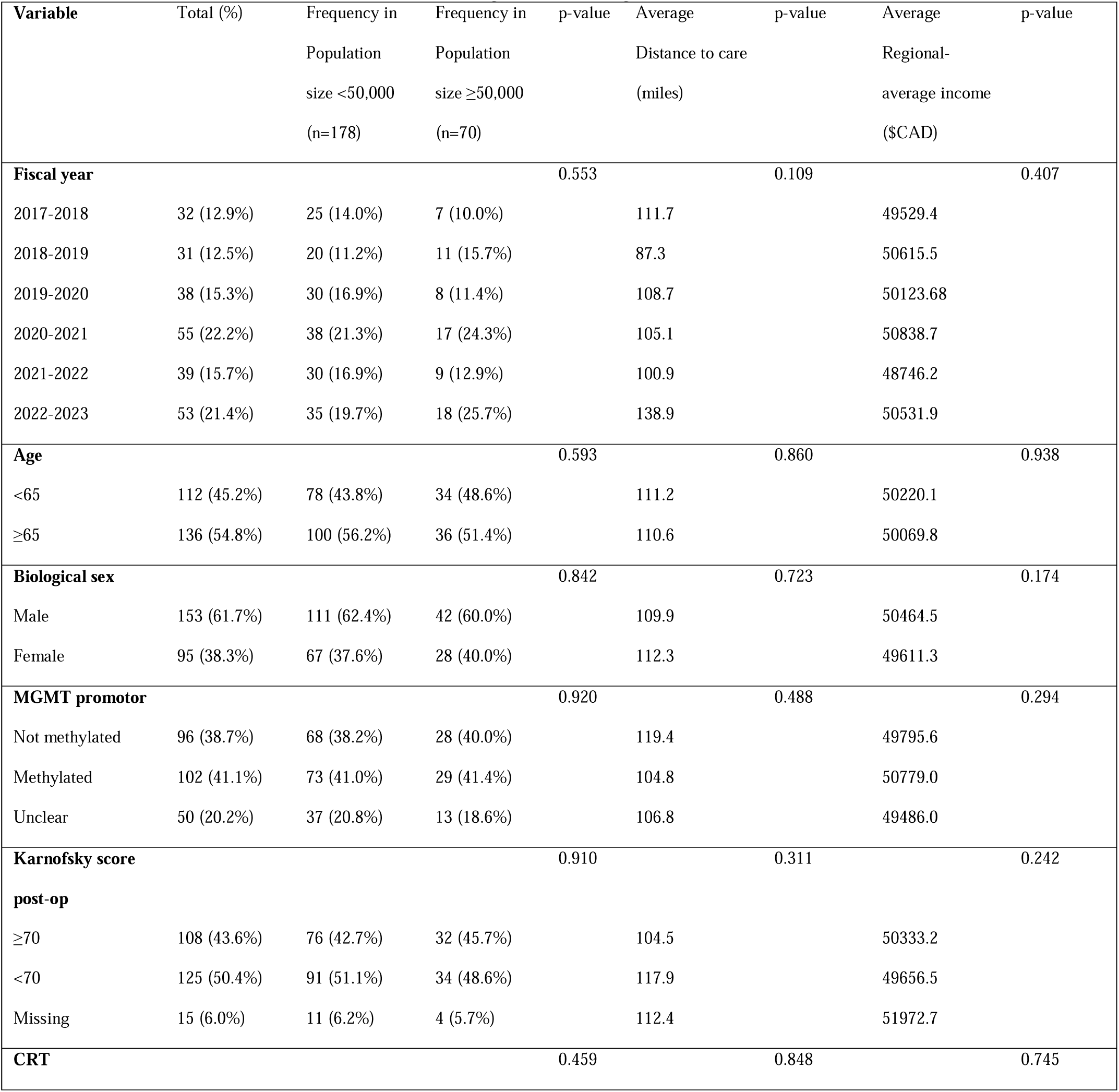
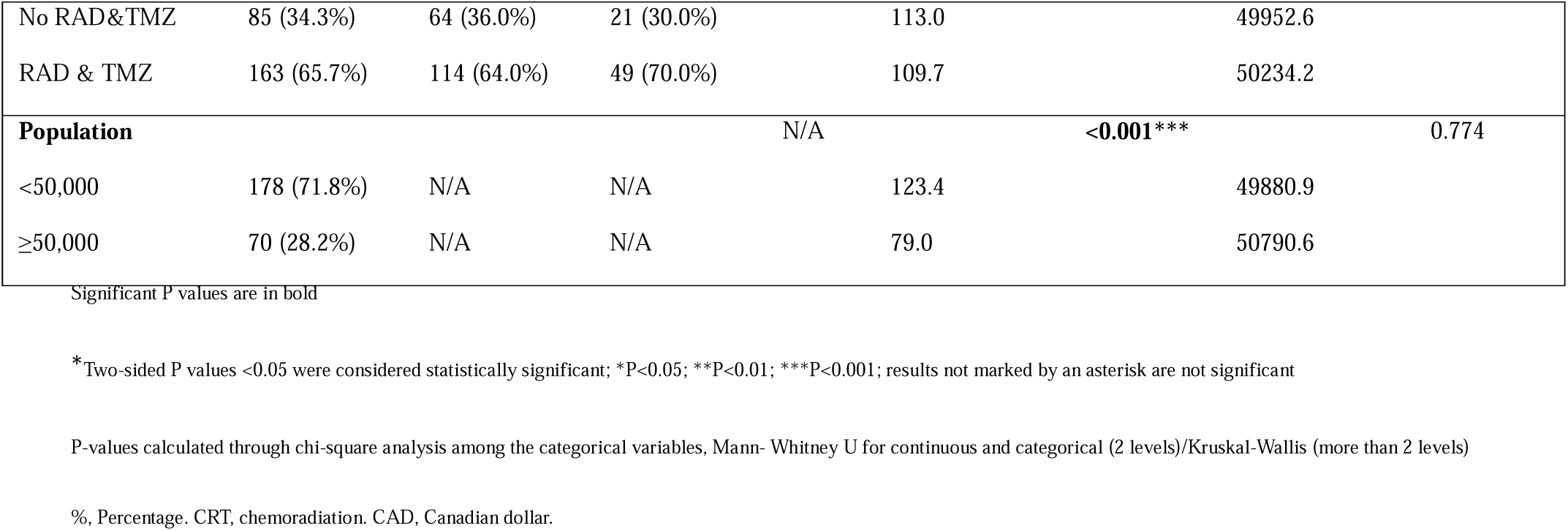
Sociodemographic, Clinical, and Geographic Characteristics of the Cohort by Population Size, Distance to Care, and Regional Average Income.

Patients originated from 87 subdivisions across SEO. Five subdivisions had populations ≥50,000 residents, while 82 had <50,000 residents. The majority of patients (n=178, 71.8%) resided in smaller communities (<50,000 residents). Distance from KHSC ranged from 2 to 300 miles for larger communities (≥50,000 residents) and 17 to 391 miles for smaller communities (<50,000 residents), with an overall mean distance to tertiary care of 111 miles (median 99 miles, IQR: 65.5-157.5; range: 2-391 miles). Patients from smaller communities had significantly greater average distance to tertiary care. Regional average income demonstrated a mean of $50,138 CAD (median $50,400 CAD, IQR: $46,960-$53,750 CAD; range: $29,600-$63,000 CAD). No significant differences in categorical confounders were observed across levels of population or regional average income (Table 1).

Mean follow-up duration was 334 days (median 219.5 days). Restricted mean OS was 12.7 months, with an overall 90-day mortality rate of 27.8% (Supplementary Figure 1). Among the 22 survivors (8.9%) at end of follow-up, mean OS was 971 days (approximately 2.7 years).

### Association of clinical and sociodemographic determinants on 90-day mortality

Of the sociodemographic variables investigated, population size (OR=0.44 [95% CI: 0.22-0.88]) and distance to tertiary care (OR=1.04 [95% CI: 1.00-1.08], per 10 miles) were associated with differences in dying within 90-days in the univariable logistic regression model (Figure 2). Notably, patients residing in areas with < 50,000 residents had a 32.0% 90-day mortality rate compared to only 17.1% of patients from areas with ≥ 50,000 residents (Supplementary Table 1). Regional average-income was not significant (OR=0.85 [95% CI: 0.50-1.47], per $10,000CAD).

**FIGURE 2.**
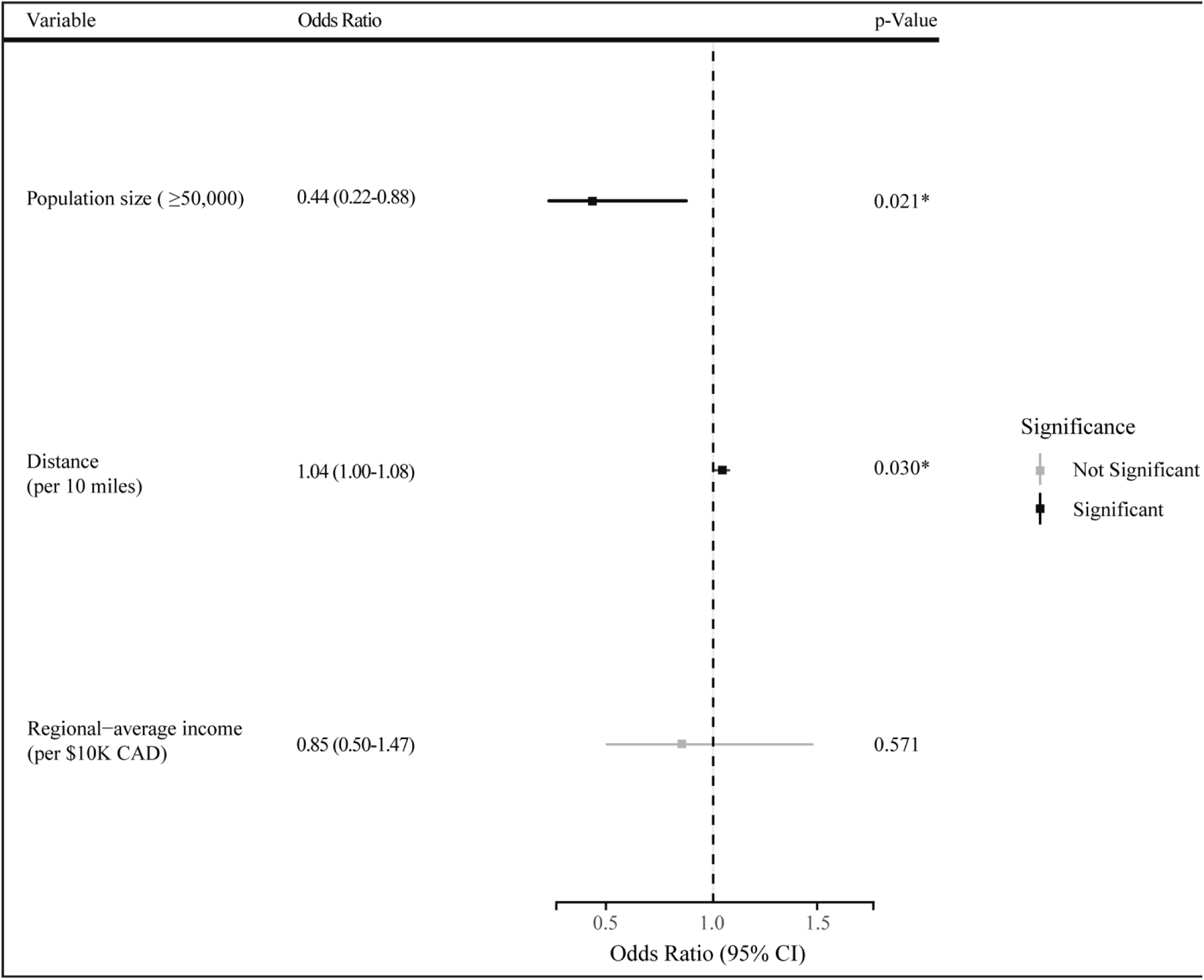
Unadjusted Odds Ratio for 90-day Mortality of Distance to Tertiary Care, Population Size, and Regional-Average Income.

In the multivariable analysis, patients residing in areas with ≥ 50,000 residents were associated with 63% *lower* odds (OR=0.37 [95% CI: 0.17-0.81]) of dying within 90-days compared to patients from less populated areas (Table 2). Meanwhile, each 10 miles of increased distance from tertiary care was associated with 5% increased odds of dying within 90-days in the multivariable analysis (OR=1.05 [95% CI: 1.01–1.10]). Regional-average income (OR=0.96 [95% CI: 0.51–1.81]) was not associated with differences in 90-day mortality in the multivariable model.

**Table 2.**
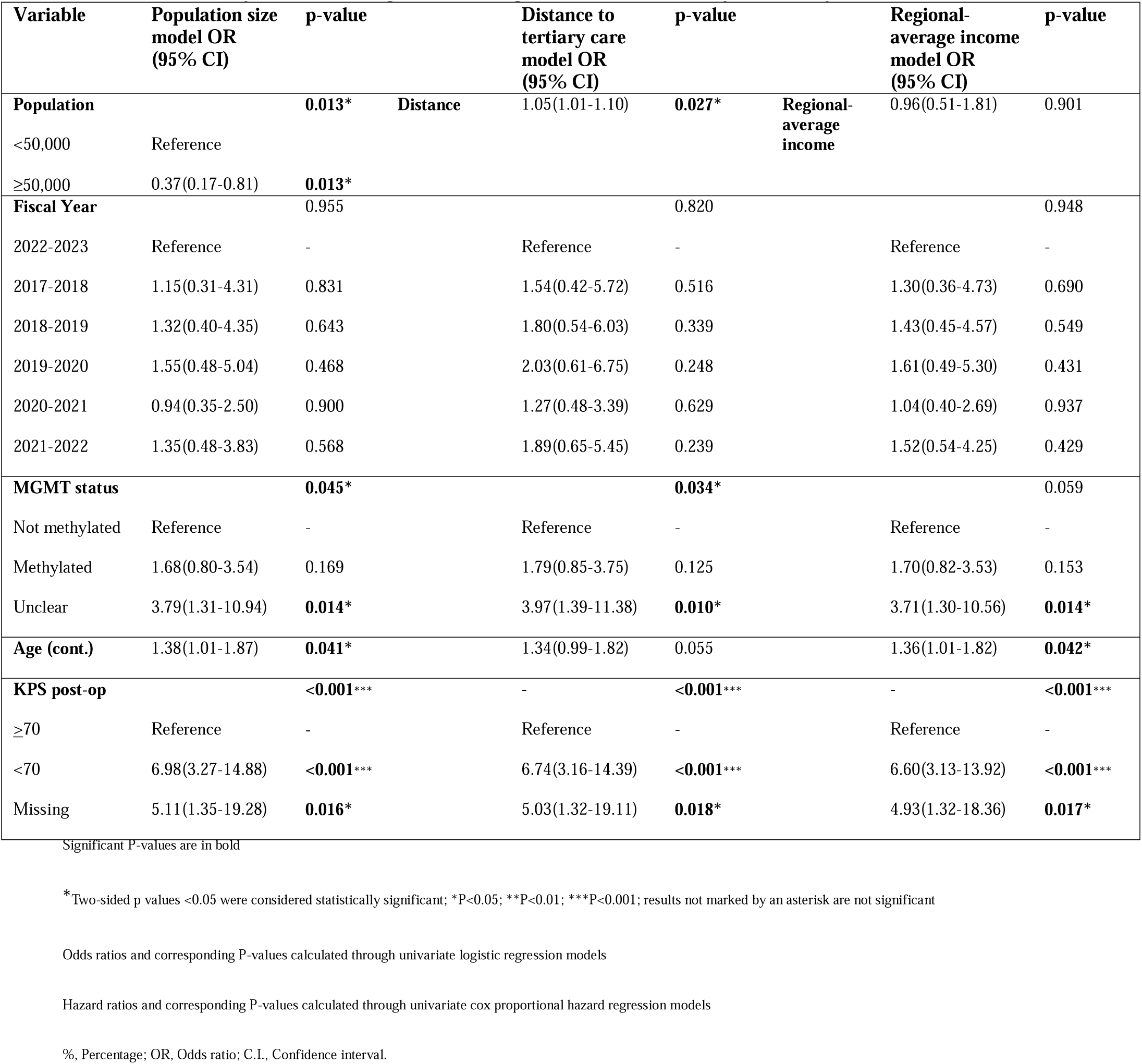
Multivariable logistic regression models examining the association of population size, distance to tertiary care, and regional average income for 90-day mortality.

Given the significant associations between population size, distance to tertiary care and 90-day mortality in their respective adjusted logistic regression models, we conducted a *post-hoc* test of interaction to explore whether these variables modified each other’s effects on the outcome. Distance effect was not significant when population size was <50,000 (p=0.114) or when population size was ≥50,000 (p=0.482), indicating no evidence that distance influences 90-day mortality differently across population sizes (Figure 3).

**Figure 3.**
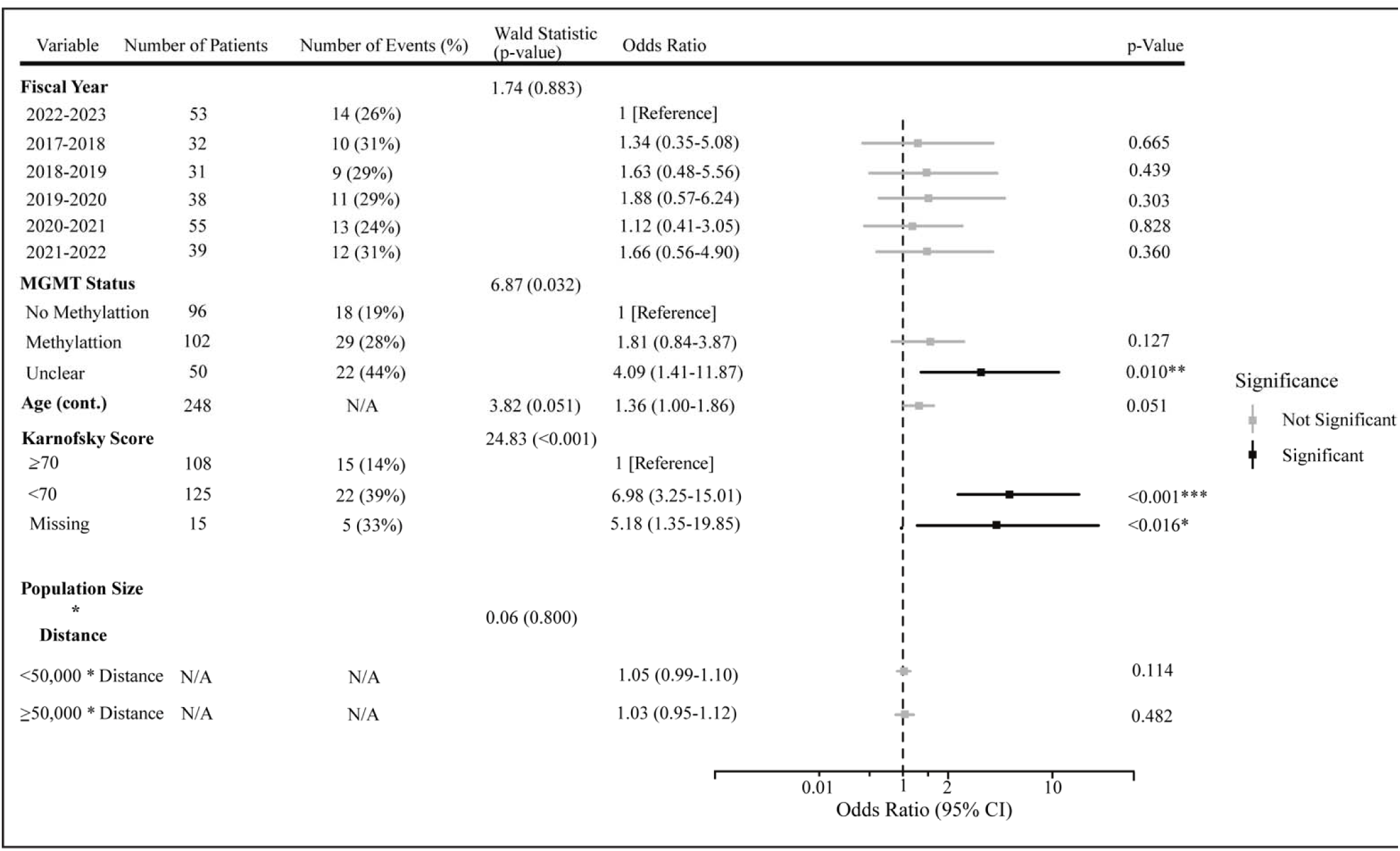
Full Adjusted Odds Ratios and 90-day Mortality Rates for all Variables Within the Interaction Model, Showing the Effects of Distance Within Each Population Size Category.

### Association of clinical and sociodemographic variables on overall mortality hazard

In the univariable Cox proportional hazard analysis, population size, distance to tertiary care and regional average income had no significant changes association with overall mortality hazard in their respective models (Supplementary Table 1). In each of the three multivariable Cox proportional hazard models, the additional variables included: fiscal year of diagnosis, sex, age (presented as per 10 years), the time varying variables (RAD and CTX), along with either population size, distance to tertiary care (presented as per 10 miles), or regional-average income (presented as per $10,000 CAD). To satisfy the proportional hazards assumption, MGMT status and KPS were included as stratification terms in the Cox model, allowing for different baseline hazards across their levels. Among the three sociodemographic variables of interest; population size, distance to tertiary care, and regional average income, none were significant in their respective multivariable Cox proportional hazard models. Living in more populated areas was associated with a 24% reduction in mortality hazard, however this did not reach significance (HR=0.76 [0.55-1.05], p=0.099). Distance to tertiary care centre (HR=1.00 [0.98-1.02]), and regional-average income (HR=0.95 [0.71-1.27]) likewise demonstrated no significant associations (Table 3). Restricted mean OS was approximately 4 months longer for patients residing in areas with ≥50,000 residents compared to those in areas with <50,000 residents, with a median OS survival advantage of approximately 3 months (Supplementary Figure 2), though this did not reach significance (p=0.100). *Post-hoc* interaction analyses between significant sociodemographic variables were not warranted, as none of the sociodemographic variables reached significance.

**Table 3.**
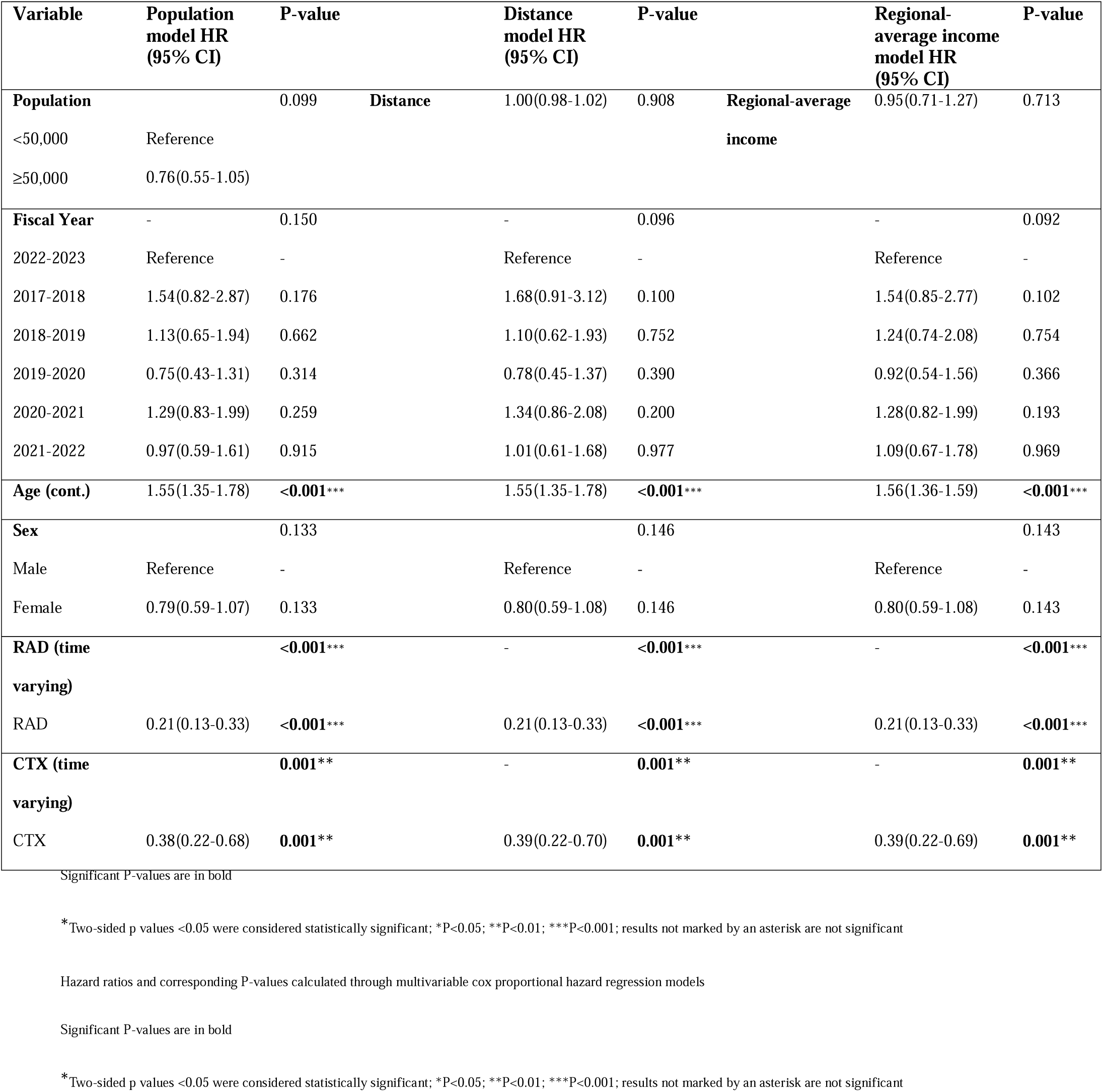
Multivariable Cox Proportional Hazards Models Examining the Association of Population Size, Distance to Tertiary Care, and Regional Average Income for Overall Mortality Hazard.

## Discussion

In a geographically disperse cohort where all patients received standardized treatment at a single tertiary center, we found that population size and geographic distance were associated with early post-operative mortality but not with OS. Patients from communities with fewer than 50,000 residents had 63% higher odds of 90-day mortality (OR=0.37 [95% CI: 0.17-0.81]), and each 10-mile increase in distance conferred 5% higher odds (OR=1.05 [95% CI: 1.01-1.10]). However, neither variable significantly affected overall mortality hazard in multivariable models. Regional average income showed no association with either outcome (90-day mortality OR=0.96 [95% CI: 0.51-1.81], p=0.901). These findings suggest that when treatment practices are standardized, rural disadvantage in GB care manifests primarily during the critical early post-operative period rather than influencing long-term outcomes.

Our findings provide important context to the mixed literature on rurality and GB outcomes. Several NCDB-based studies have reported survival disadvantages for patients residing in lower populated areas, with Ostrom et al (2021) showing that urban patients had better OS than rural patients (HR=0.93 [95% CI: 0.86-1.00])^24^ and Estevez-Ordonez et al (2023) reporting an even stronger rural disadvantage (HR=1.79 [95% CI: 1.17–2.73])^12^. However, other studies have found no significant association between population size and overall mortality hazard^7,46^. These inconsistencies may reflect methodological limitations. Multi-center databases often underrepresent patients from lower-populated areas and risk confounding population density with treatment access. For example, Hodges et al. (2021) reported no significant difference in 90-day mortality between rural and metropolitan patients in their NCDB cohort, but rural patients comprised only 2% of their sample despite 19% of the US population residing in rural areas^5^, raising concerns about representativeness. In keeping, Ramapriyan et al (2023) demonstrated that rural patients have substantially reduced access to RAD (OR=0.34 [95%CI: 0.25-0.47]) and CTX (OR=0.42 [95%CI: 0.34-0.52])^26^, suggesting that survival differences in multi-center studies may reflect disparities in treatment delivery rather than independent effects of population density. Our results from a cohort with 71.8% rural representation and standardized treatment practices suggest a potential resolution. When treatment access within the tertiary care center is equalized, the primary impact of population size manifests in the immediate post-operative period rather than affecting long-term mortality hazard.

Our finding that greater distance from tertiary care was associated with increased 90-day mortality contrasts with paradoxical findings in prior studies, where longer travel distances were associated with improved survival outcomes^28,47^. These counterintuitive results may reflect selection bias in multi-center databases: patients who travel farther in NCDB cohorts may be more likely to receive care at higher-volume or private facilities, representing a self-selected population with greater resources and health-seeking behavior. In contrast, all patients in our study were treated at the same tertiary care center within a publicly funded healthcare system, eliminating variability in institutional quality. Bird et al. provided evidence for this selection mechanism, demonstrating that while travel distances over 50 miles were associated with improved OS, this protective effect was driven entirely by patients in the highest income quartile (>$63,000 USD)^6^, suggesting that only individuals with substantial financial resources can overcome geographic barriers to access specialized care. Our finding of increased 90-day mortality with distance, observed in a cohort where institutional factors are held constant, suggests that when selection bias is reduced, geographic distance imposes barriers to post-operative care.

The concentration of rural disadvantage in the early post-operative period likely reflects challenges unique to this 90-day window. This period is characterized by important transitions including hospital discharge, wound healing, complication management, and initiation of adjuvant therapy. Patients from lower-populated areas may face greater barriers during this vulnerable period due to reduced access to specialized post-operative care, limited availability of rehabilitation services, and decreased social support networks in sparsely populated communities ^48^. Greater distance from tertiary care may create challenges attending follow-up appointments, delays in identifying and managing post-operative complications, difficulty coordinating care transitions between tertiary and local providers, and lack of neurosurgical expertise at regional emergency departments for acute post-operative complications. These geographic and structural barriers appear to exert their greatest impact when medical needs are most acute, and care coordination is most essential.

Our finding that distance and population size exert independent effects on 90-day mortality, with no significant interaction between them, has important implications for health system interventions aimed at reducing rural disparities. This independence suggests that simply bringing care closer to rural communities through “closer to home” initiatives may be necessary but insufficient to eliminate outcome disparities. While such initiatives address geographic barriers^49^, they do not resolve challenges inherent to low population density, including limited local healthcare infrastructure, reduced availability of specialized rehabilitation and home care services, and smaller social support networks. Our results indicate that patients from sparsely populated areas require comprehensive interventions beyond geographic proximity alone. The first 90 days post-surgery may benefit from enhanced care coordination models that include proactive telehealth follow-up, partnership with local providers for complication surveillance, dedicated patient navigators to facilitate care transitions, and community-based support programs tailored to rural populations. By recognizing that distance and population density operate through distinct mechanisms, health systems can design targeted interventions that address both geographic and structural barriers to equitable post-operative care.

Our finding that regional average income was not associated with either 90-day mortality or overall mortality hazard contrasts with several studies reporting protective effects of higher income on GB outcomes^3,7,15–17^. This discrepancy likely reflects fundamental differences in healthcare system structure rather than methodological variation. Studies demonstrating income-related survival advantages have predominantly analyzed large databases from private care systems ^3,7,16,17^, where socioeconomic status impacts access to timely diagnosis as well as specialist consultation and therapies. In contrast, studies conducted within public healthcare systems^22^ or at single tertiary centers^18–21,23^, such as ours, consistently report null effects of income on survival. This pattern suggests that universal healthcare coverage may attenuate income-related disparities in treatment access. Our results therefore suggest that socioeconomic disparities in GB outcomes may be modifiable through system-level policies rather than reflecting inevitable consequences of income inequality.

Our findings should be interpreted in light of several limitations. First, our sample size of 248 patients may have limited statistical power to detect a significant association between population size and overall mortality hazard, though the observed HR of 0.76 suggests a trend toward improved survival in larger populations. Second, approximately 20% of our cohort had unclear MGMT methylation status and 6% had missing KPS data. Third, as a single-center retrospective study, generalizability may be limited to regions with similar healthcare delivery models and geographic characteristics. Finally, we were limited to using area-level income as a proxy for individual-level income. While Canadian studies have shown similar associations between individual- and area-level income for health outcomes, there is poor agreement in how patients are classified. For example, in a study of 103,530 colorectal cancer patients, only 17% of individuals fell into the same income quartile when comparing individual- and neighborhood-level measures^50^. This misclassification is substantially worse in rural areas, suggesting that census-subdivision income may poorly approximate the true impact of individual income on GB outcomes in our predominantly rural cohort.

These findings highlight critical opportunities for intervention during the early post-operative period to reduce rural disparities in GB outcomes. Targeted care coordination models that address the distinct barriers of distance and low population density may mitigate the disadvantages observed in the first 90 days after surgery. Further research is needed to evaluate whether such interventions can eliminate rural-urban outcome gaps in GB care.

## Supporting information

Supplementary Table 1, Supplementary Figures 1&2, and Supplementary Section 1

## Required Statements

### Ethical Consideration

Ethical approval was obtained from the Queen’s University Health Sciences Research Ethics Board (HSREB; TRAQ# 6043517).

## Contributions

M.L, A.Ma, S.T, K.G, and T.P contributed to the study design. Data collection was conducted by M.L, A.Mo, D.T, S.A, K.A, and T.P. Statistical analyses were performed by M.L, S.T, and J.P. The manuscript was written by M. L, A.Ma, S.T, K.G, and T.P. All authors reviewed and approved the final manuscript.

## Data Availability

Deidentified patient data supporting this study are available from the corresponding authors upon reasonable request, subject to institutional and ethics approval.

## Use of Artificial Intelligence Disclosure

During the preparation of this manuscript, the authors used large language model chatbots such as ChatGPT for language editing including grammar, readability, and sentence conciseness. After using this tool, all authors reviewed and edited the content provided by ChatGPT to ensure the content aligned with the manuscript.

