## Supplementary Table 1, Supplementary Figures 1&2, and Supplementary Section 1 for "Rural Disadvantage in Glioblastoma Concentrates in the Early Postoperative Period: A Single-Center, Treatment-Standardized Cohort Study"

Supplementary Tables and Figures

**Supplementary TABLE 1**. Univariate Logistic Regression Models for Predicting Association Odds of 90-day Mortality and Overall Survival Among Categorical and Continuous Independent Variables.

| **Variable** | 90-day Mortality (%) | 90-day mortality OR (C.I.) | p-Value | HR for OS (C.I.) | p-value |
| --- | --- | --- | --- | --- | --- |
| **Fiscal year**  2022-2023  2017-2018  2018-2019  2019-2020  2020-2021  2021-2022 | 14 (26.4)  10 (31.3)  9 (29.0)  11 (28.9)  13 (23.7)  12 (30.8) | Ref  1.27 (0.48,3.32)  1.14 (0.42,3.06)  1.13(0.45,2.88)  0.86(0.36,2.06)  1.24(0.50,3.09) | -  0.632  0.795  0.790  0.739  0.647 | Ref  1.23(0.77,1.95)  0.87(0.54,1.39)  0.79(0.50,1.24)  0.84(0.56,1.27)  1.03(0.66,1.61) | -  0.385  0.558  0.300  0.407  0.884 |
| **Pathology**  GB  Other | 68 (28.8)  1 (8.3) | Ref  0.22(0.03,1.77) | -  0.157 | Ref  0.55(0.29,1.05) | -  0.070 |
| **MGMT promotor**  Not methylated  Methylated  Unclear | 18 (18.8)  29 (28.4)  22 (44.4) | Ref  1.72(0.88,3.36)  **3.40(1.60,7.27)** | -  0.112  **0.002**** | Ref  0.78(0.58,1.05)  1.28(0.90,1.83) | -  0.107  0.171 |
| **Age**  ≥65  <65 | 21 (18.8)  48 (35.3) | Ref  **0.42(0.23,0.76)** | -  **0.004**** | Ref  **0.45(0.34,0.59)** | -  **<0.001***** |
| **Age (cont.)** | **N/A** | **1.61(1.23,1.76)** | **<0.001***** | **1.56(1.37,1.78)** | **<0.001***** |
| **Biological sex**  Male  Female | 43 (28.1)  26 (27.4) | Ref  0.96(0.54,1.71) | -  0.900 | Ref  0.85(0.65,1.12) | -  0.249 |
| **Karnofsky score post-op**  ≥70  <70  Missing | 15 (12.0)  49 (45.4)  5 (33.3) | Ref  **6.09(3.15,11.77)**  **3.67(1.60,12.19)** | -  **<0.001*****  **0.034*** | Ref  **2.12(1.61,2.80)**  1.41(0.81,2.46) | -  **<0.001*****  0.226 |
| **RAD flag** |  | N/A | **-** | **0.25(0.17,0.34)** | **<0.001***** |
| **CTX flag** |  | N/A | **-** | **0.33(0.22,0.52)** | **<0.001***** |
| **Population**  <50,000  ≥50,000 | 57 (32.0%)  12 (17.1%) | Ref  **0.44 (0.22,0.88)** | -  **0.021*** | Ref  0.78(0.58,1.05) | -  0.099 |
| **Distance(cont.)** |  | **1.04(1.00,1.08)** | **0.030*** | 1.01(0.99,1.03) | 0.210 |
| **Average income area(cont.)** |  | 0.85(0.50,1.47) | 0.571 | 1.00(0.77,1.29) | 0.985 |

^Significant P-values are in bold^

**^*^**^Two-sided p values <0.05 were considered statistically significant; *P<0.05; **P<0.01; ***P<0.001; results not marked by an asterisk are not significant^

^Odds ratios and corresponding P-values calculated through univariate logistic regression models^

^Hazard ratios and corresponding P-values calculated through univariate cox proportional hazard regression models^

^%, Percentage; OR, Odds ratio; C.I., Confidence interval; OS, Overall survival; HR, Hazard ratio; RAD, Radiation Therapy; CTX, Chemotherapy^

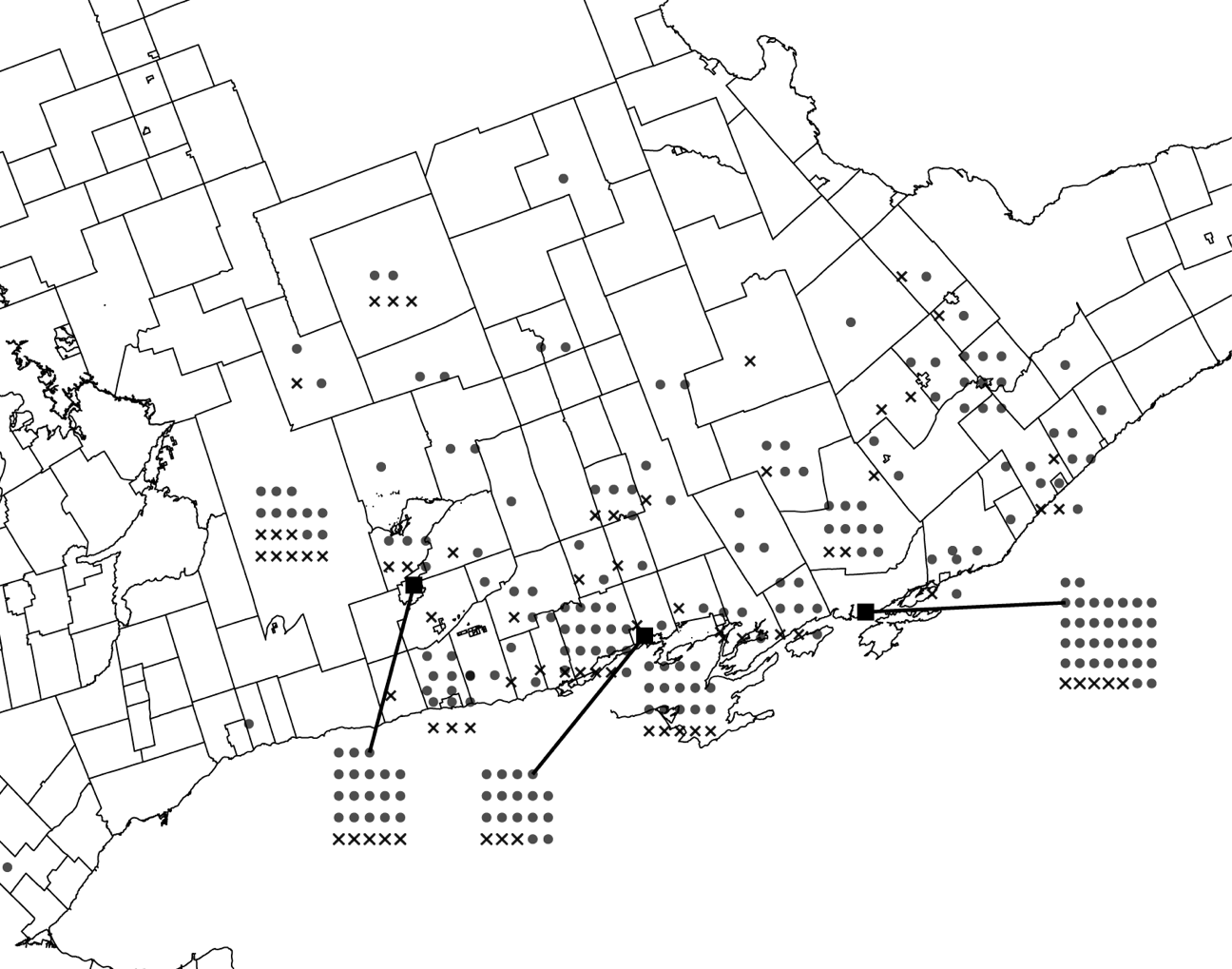

**Supplementary Figure 1.** Geographic distribution of 90-day mortality across census subdivisions in Southeastern Ontario. Bolded squares indicate the cities of Peterborough (left), Belleville (center), and Kingston (right).

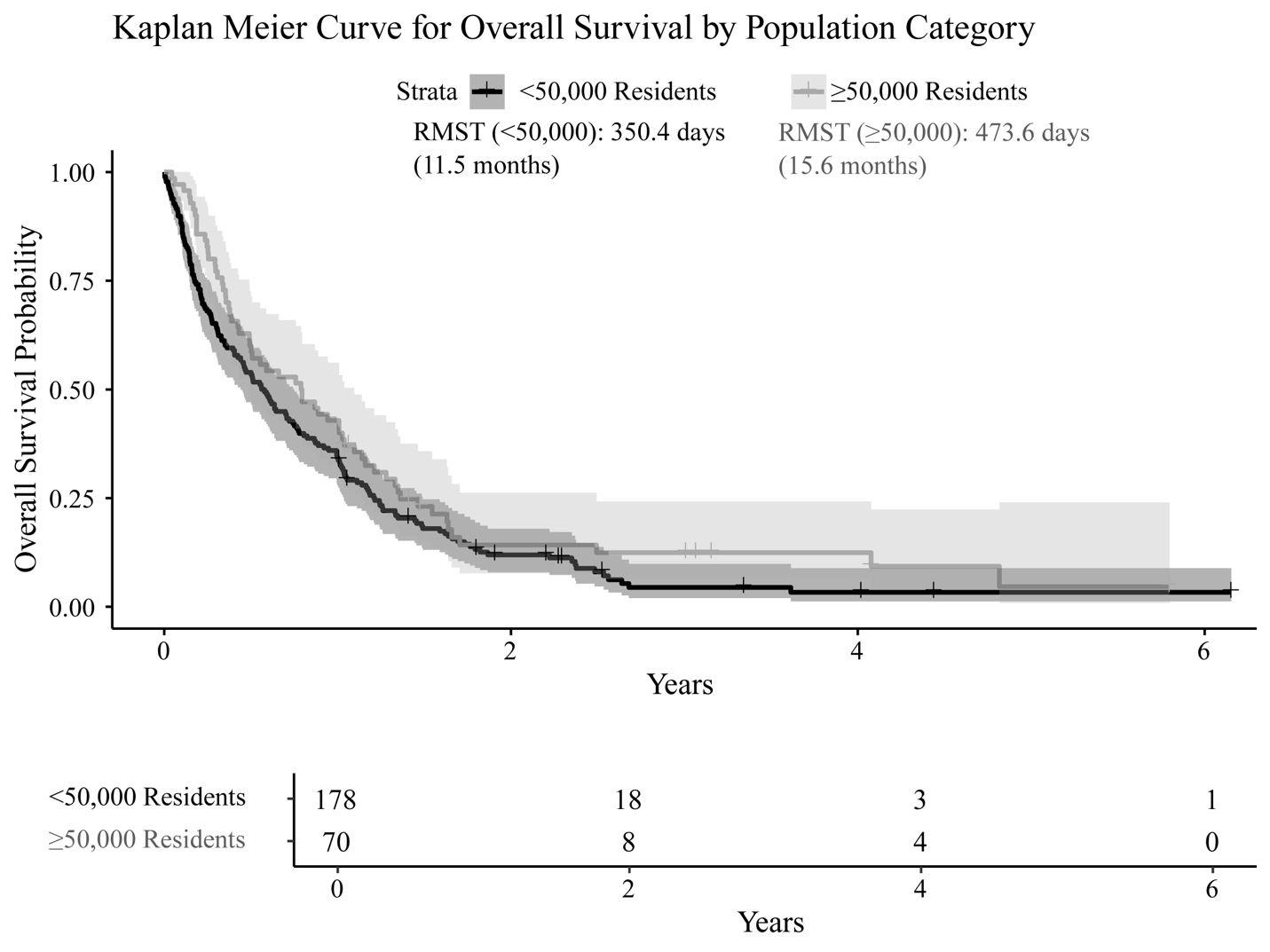

**Supplementary Figure 2.** Kaplan–Meier Curve for Overall Survival Estimating Overall Survival Probability Over Time (in years) By Population Categories. Shaded areas represent 95% confidence intervals.

^Restricted Mean Survival Time (RMST) refers to the average survival time up to the end of follow-up, calculated separately for each population category.^

**Supplementary Section 1**:

While age and KPS are established risk factors associated with poorer survival outcomes, “unclear MGMT methylation status” also consistently emerged as a significant risk factor for 90-day mortality. In our center, MGMT status is evaluated externally and requires closer to 30 days to receive. If there is insufficient tissue, or if the patient is deceased prior to the receipt of MGMT status, then MGMT status is deemed “unclear”. “Unclear MGMT status” may be a significant predictor of worse outcomes because it reflects an enrichment of patients with early mortality, or those whose tumors were not amenable to resection, resulting in insufficient tissue profiling. Unsurprisingly, receipt of RAD and CTX were both highly significant predictors of OS, even when modeled as time-varying variables.
